# Use of Physician Global Assessment (PGA) in Systemic lupus erythematosus: a systematic review of its psychometric properties

**DOI:** 10.1101/2020.04.14.20064683

**Authors:** Elisabetta Chessa, Matteo Piga, Alberto Floris, Hervé Devilliers, Alberto Cauli, Laurent Arnaud

**Affiliations:** Rheumatology Unit, AOU University Clinic and University of Cagliari, Cagliari, Italy; Centre Hospitalier Universitaire de Dijon, Hôpital François-Mitterrand, service de médecine interne et maladies systémiques (médecine interne 2) et Centre d’Investigation Clinique, Inserm CIC-EC 1432, Dijon, France; Service de rhumatologie, Ho□pitaux Universitaires de Strasbourg, Université de Strasbourg, Strasbourg, France; Centre National de Référence des Maladies Systémiques et Autoimmunes Rares Est Sud-Ouest (RESO), Strasbourg, France

## Abstract

**Background:** Physician Global Assessment (PGA) is a visual analogue score (VAS) that reflects the clinician’s judgment of overall Systemic Lupus Erythematosus (SLE) disease activity. The aim of this systematic literature review (SLR) is to describe and analyse the psychometric properties of PGA.

**Methods:** This SLR was conducted by two independent reviewers in accordance with the PRISMA statement. All articles published until the 1st of July 2019 in Pubmed were screened with no limitation about years of publication, language or patients’ age. Psychometric properties data were analysed according to the OMERACT Filter methodology version 2.1.

**Results:** The literature search identified 91 studies. Face validity was reported in all the articles retrieved, in which the PGA was used alone or as part of composite indices (SRI, SFI, LLDAS, DORIS remission criteria). Content validity was reported in 89 studies. Construct validity was demonstrated by a good correlation (r≥0.50) between the PGA with the SLEDAI (12 studies), SLAM (4 studies), LAI, BILAG and ECLAM (2 studies each). Criterion validity was assessed exploring the PGA correlation with quality of life measurements, biomarkers levels and treatment changes in 28 studies, while no study has evaluated correlation with damage. A good responsiveness for PGA was shown in 8 studies. A high variability in scales was found, causing a wide range of reliability (ICC=0.67-0.98).

**Conclusion:** PGA is a valid, responsive and feasible instrument, while its reliability was impacted by the scale adopted, suggesting the major need for a standardization of its scoring.

## 1. Introduction

Systemic lupus erythematosus (SLE) is an immune-mediated multi-systemic disease characterised by a wide variability of clinical manifestations and a course frequently subject to unpredictable flares (1-2). Because of SLE complexity, the assessment of disease activity is particularly challenging (3). The judgment of whether a patient with SLE has active disease is a central question, both in routine patient management as well as in clinical research (4). A higher disease activity is an important predictor of both organ damage and mortality (2) and the attainment of at least a low disease activity is associated with a reduction of early damage (5-6).

In the last 30 years more than a dozen of scores have been derived to assess disease activity in SLE, but not all of these have proven valid and reliable tools. Nevertheless, the development of a comprehensive index for assessing disease activity still represents one of the most important challenges in SLE (7).

In 1988, Liang et al. (8) suggested the need for a disease activity instrument which would be valid, reliable and sensitive to change. They proposed a physician global scoring on a 10-cm visual analog scale (VAS) to be used as a gold standard to compare six disease activity instruments (4). The term “Physician Global Assessment” (PGA) was coined in 1991 by Petri et al (9) to address a disease activity index scored on a VAS ranging from 0 to 3, with an increase of ≥1.0 since the last visit indicating a flare. Subsequently, the PGA was incorporated in the SELENA flare index in 1999 (10), in the Systemic Responder Index in 2009 (3, 11-12) as well as in the definitions of low disease activity (LLDAS) (13) and various definitions of remission (14-15).

The last EULAR/ACR recommendations recommend the use of the PGA in the routine monitoring of SLE (16). However, no precise guidelines exist regarding the optimal use of the PGA in SLE, such as the adequate length of the VAS, the presence of anchored values, the incorporation of laboratory data and the time frame of assessment.

The aim of this systematic review is to describe and analyse the measurement properties of the PGA, including the validity, reliability, responsiveness and feasibility.

## 2. Material & Methods

### 2.1. Literature search

A systematic review of the literature was performed according to the PRISMA guidelines (17), searching for articles reporting on the use of PGA in SLE. The following search strategy was used through MEDLINE via PubMed: (((“lupus erythematosus, systemic”[MeSH Terms] OR (“lupus”[TIAB] AND “erythematosus”[TIAB] AND “systemic”[TIAB]) OR “systemic lupus erythematosus”[TIAB] OR (“systemic”[TIAB] AND “lupus”[TIAB] AND “erythematosus”[TIAB]))) OR “SLE”[TIAB]) AND (“physician global assessment”[TIAB] OR “PGA”[TIAB]). Additional papers were obtained by checking the references from the selected studies. Reviews and case series with less than 5 patients were excluded. Retrieved papers were selected with no limitation about years of publication, language or patients’ age. The last Medline search was performed on the 1st of July 2019. Once 2 investigators (E.C., M.P.) have independently selected the articles, initially on the basis of titles and abstracts then, if necessary, on the full texts, eligibility assessment was performed independently in a blinded standardized manner. Disagreements between investigators were solved by consensus. Whenever papers reported duplicate data, the most recent article was selected.

### 2.2 Extraction of Psychometric Properties of the PGA

Each study was examined in order to extract psychometric property data on PGA according to the OMERACT (Outcome Measures in Rheumatology) Filter methodology version 2.1 (18). Measurement properties of the PGA were analysed according to the COnsensus-based Standards for the selection of health Measurement INstruments (COSMIN) terminology (19).

## 3. Results

### 3.1. Literature search

The literature search identified 93 articles, and 12 additional articles were retrieved from the reference list of those publications. A total of 91 articles were included in the study (Fig. 1), accounting for 49 longitudinal cohort studies, 25 cross-sectional studies, 7 Randomized Controlled Trials (RCT), 3 consensus conferences, 4 post-hoc analyses, 2 retrospective studies and 1 case-series.

**Fig. 1:**
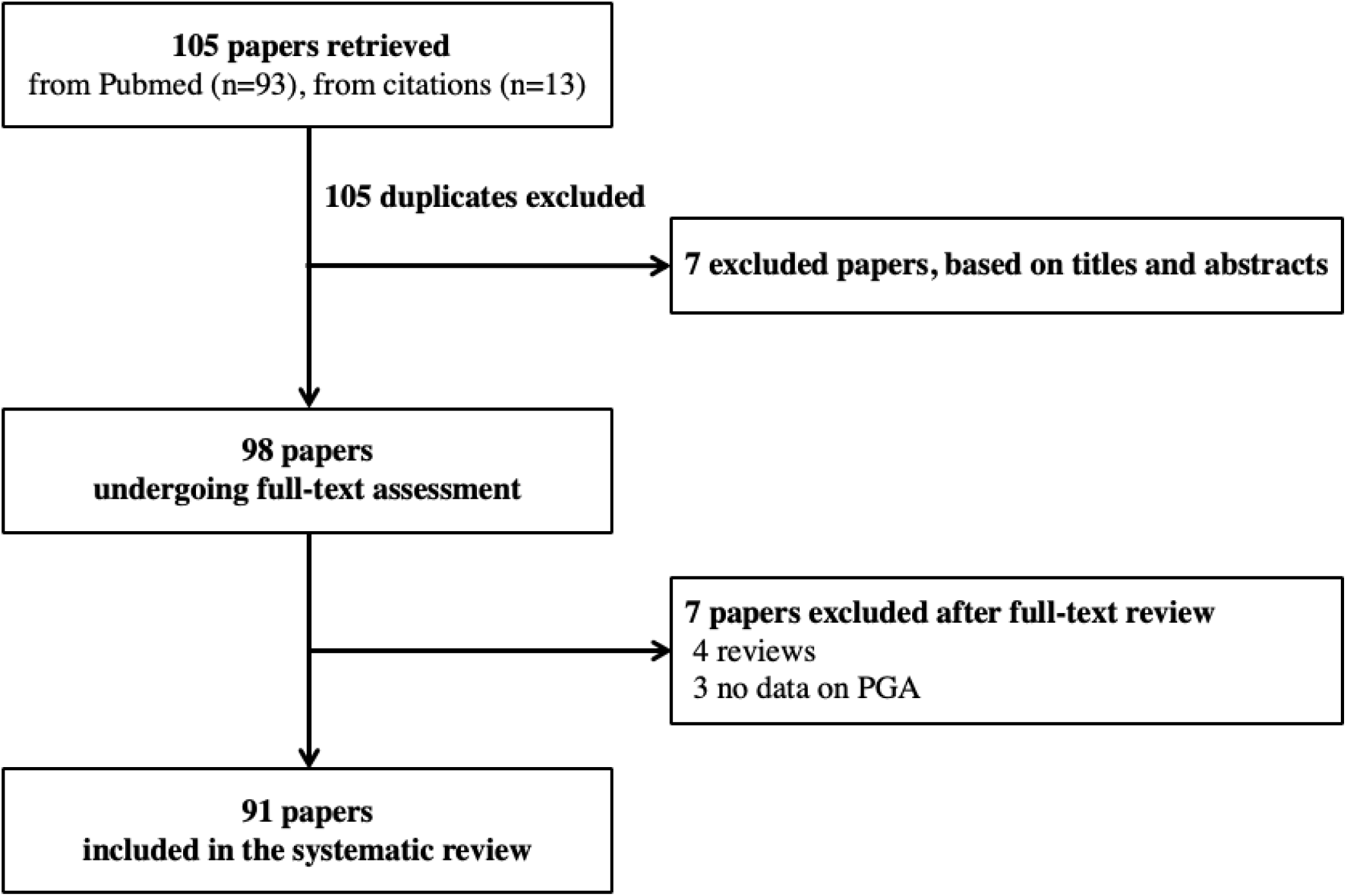
Flow chart illustrating the literature search and study selection.

### 3.2 Psychometric Properties of the PGA

The OMERACT defines an instrument fit as outcome measurement if it passes the three pillars of evidence: truth (that refers to validity), discrimination (that includes reliability and responsiveness) and feasibility.

#### 3.2.1 Truth (validity)

Truth refers to whether the measure provided by the scores is able to measure what was intended for (18). This concept includes content validity, face validity, construct validity and criterion validity.

##### Content validity

Content validity pertains to the degree to which the instrument measures all facets of a construct of interest (20): this property is satisfied if the PGA is considered able to measure all aspects of disease activity in SLE, in a comprehensive way. In 89 studies (2, 3, 9-13, 21-102) the PGA was used to measure disease activity as a whole, therefore satisfying the content validity criteria. The globality of measurement was intended in the form of a scale from 0 to 3 in 54 studies (2, 3, 9-10, 12-13, 21, 24–69, 103) in a 0-10 scale in 12 studies (4, 60, 70-79), 0-100 in 9 studies (27, 40, 78, 80-85), 0-7 Likert scale (11, 78, 80), 0-2 scale (53), 0-4 (86) and 0-5 (87). Different definitions of PGA retrieved through literature search are reported in table 1.

**Table 1:** Different definitions of disease activity according to PGA instrument.

| **Disease activity status** | **PGA values**<br>**(scale)** | **PGA**<br>**Definition** | **Study**<br>**(reference)** |
| --- | --- | --- | --- |
| **Remission** | 0-3 VAS | 0.5 | (29, 37) |
|  |  | 0 | (36, 72) |
| **Low disease activity** | 0-3 VAS | ≤1 (LLDAS) | (12-13, 29, 34, 37, 39-40) |
|  |  | <1.5 | (63) |
| **Inactive disease** | 0-3 VAS | 0 | (3, 24, 26-27, 38, 41-42, 51, 54, 60, 62, 65, 87, 99) |
|  | 0-10 VAS | 0 | (72-75, 83) |
|  | 0-100 VAS | 0 | (80, 82, 94) |
|  | 0-2 VAS | 0 | (88) |
| **Mild activity** | 0-3 VAS | 1 | (24, 26-27, 41-42, 60, 62, 65, 99) |
|  |  | <1.0 | (2) |
|  | 0-2 VAS | 1 | (88) |
| **Moderate activity** | 0-3 VAS | 2 | (24, 26-27, 42, 60, 62, 65, 99) |
|  |  | 1.1 – 2.0 | (2) |
|  |  | 2 – 2.5 | (3) |
| **Severe activity** | 0-3 VAS | 3 | (3, 24, 26-27, 41-42, 60, 62, 65, 99) |
|  |  | 2.1 – 3.0 | (2) |
|  | 0-2 VAS | 2 | (88) |
| **Flare** | 0-3 VAS | Increase of ≥1.0 |  |
|  |  | - Last 93 days | (9) |
| **Mild-moderate flare (SFI)** | 0-3 VAS | - From last visit | (10) |
| **Severe flare (SFI)** | 0-3 VAS | Increase to >2.5 | (10) |
| **Clinical worsening** | 0-3 VAS | Increase of ≥0.3 | (3) |

In the second column the definitions were reported according to the VAS used in the study. VAS: Visual Analogue Scale. SFI: Selena Flare Index.

In 2 studies, the PGA-VAS was used to assess concepts other than disease activity: disease severity (4) and patients’ compliance to treatments (103).

Few studies reported on whether serological activity should be incorporated in the PGA. Liang et al. (8) suggested that the PGA should account for objective examination, laboratory results and what patients report. Barr et al. (2, 21, 24) recommended PGA assessment prior to reviewing serological data, based only on clinical visit. This scoring modality was used for the SRI (3). In one study (25), PGA of disease activity resulted from the combination of clinical visit, laboratory markers evaluation and physician’s knowledge of the patient disease history. In the absence of a consensus, Aranow (26) found a better correlation between the SLEDAI and the PGA when the latter was assessed taking into account laboratory test results.

##### Face validity

According to the OMERACT, face validity is the degree to which the instrument appears to match with the target domain, according to experts (18). Face validity is satisfied when the instrument is considered able to capture what it should capture (i.e. disease activity). This property is reported across all articles selected through this systematic review (2-4, 9-13, 21-103).

Supporting the face validity property, PGA was defined “gold-standard” in 11 studies (2, 10-11, 21, 32, 49, 67, 76, 78, 84, 88) and in 32 it was used as the reference to which other activity scores were compared, such as the SLEDAI (4, 10, 13, 25, 27-28, 31, 33, 35-36, 41, 46-47, 50-51, 53, 62, 65, 68, 72-73, 76, 81, 96-99), BILAG (4, 27, 35-36, 46, 65, 72-73, 81, 98), SLAM (4, 72, 76, 99), LAI (68, 88), patient global assessment (ptGA) (81, 83-84) and ECLAM (35). However, it was used as a single outcome measure only in two studies (49, 100) whilst in the majority the PGA was scored together with another instrument (typically the SLEDAI) (2, 9, 11-12, 21, 24, 30, 32, 34, 37-40, 44-45, 48-49, 55, 58-59, 61, 63-64, 66-67, 74-75, 80, 82, 86, 89-95, 103).

In 16 studies the PGA was used as a mean to assess changes in disease activity after treatment (3, 12, 22, 29, 40, 42-43, 52, 54, 56-57, 80, 85, 87, 95, 98, 101). In one open-label study (43) the decrease in PGA score was considered the primary endpoint. In 32 studies, disease activity measured by PGA was compared to changes in laboratory markers, with the aim to correlate clinical and serological features (9, 21, 30, 32, 34, 37-39, 44-45, 48-49, 55, 58-61, 63-64, 66-67, 69, 71, 74-75, 82, 86, 89, 91-94).

The PGA was integrated in composite indices, including the definition of LLDAS (12-13, 29, 34, 37, 39-40) and remission (29, 37) (table1). In 11 retrieved studies (10, 13, 33, 36, 45, 48, 50, 55-56, 65, 96) the PGA was part of SELENA Flare Index (SFI) (104) and in 10 studies (3, 29, 40, 46, 52, 60, 69, 80, 94, 98) it was part of the Systemic Responder Index (SRI) (3) (both discussed in responsiveness paragraph). In an epratuzumab trial, the absence of deterioration of PGA (not >10% worsening) was one of the items to achieve BICLA (BILAG-Based Composite Lupus Assessment) response (105). In one study, PGA was part of a modified score to assess disease activity in pregnancy (SLEPDAI) (51).

##### Construct validity

Construct validity is the degree to which the PGA relates to other instruments which measure the same concept (18). This may be explored through convergent and divergent validity. Data regarding divergent validity are lacking for PGA. Convergent validity is fulfilled indirectly in studies where the PGA is used as the gold standard to assess the construct validity of other indices. Correlations with other instruments measuring similar constructs should typically demonstrate coefficient (r)≥0.50 (106).

Construct validity was recognized in 21 studies (2, 10-11, 23-24, 26-29, 35, 47, 52, 54, 65, 68, 76, 84, 88, 92, 99, 101). The correlation with the SLEDAI was determined in 12 studies (fig. 2) (10, 23-24, 26, 28-29, 35, 54, 68, 76, 84, 99), with the SLAM in 4 studies (r comprised between 0.47 and 0.65) (35, 76, 84, 99), with LAI in 2 studies (r=0.64-0.75) (68, 84), with BILAG in 2 studies (r=0.61-0.62) (35, 84) with ECLAM in 2 studies (r=0.58-0.65) (35, 84).

**Fig. 2:**
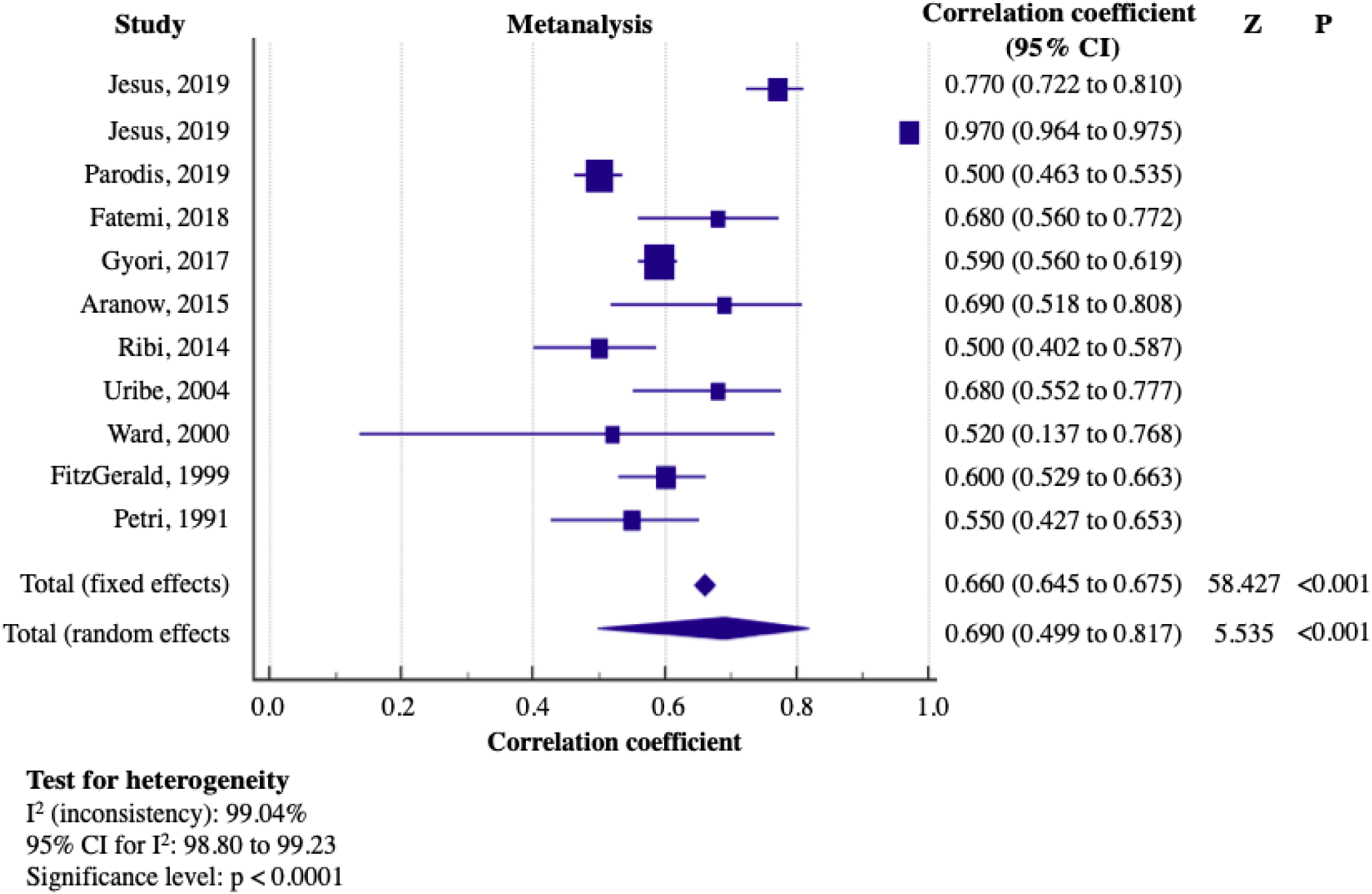
Meta-analysis of the studies reporting data concerning the construct validity between PGA and SLEDAI. A good correlation was considered for a value higher than 0.60. The pooled correlation coefficients (95% CI) is given both for the fixed effects model and the random effects model. Random effects model gives a more conservative estimate considering the heterogeneity.

##### Criterion validity

Criterion validity is defined as the degree to which the scores of an instrument adequately reflect “the truth” in the form of a “gold standard” (107). In the absence of a well-recognised gold standard for disease activity, criterion validity of PGA is established when it correlates with a measure that the author of the study defined *a priori* as the gold standard. In Fatemi et al. (35) the PGA correlated, although moderately, with the need for treatment change (r=0.46, p<0.01).

Criterion validity also refers to the degree to which an instrument predicts aspects and phenomena occurring in the future (108). In this sense, criterion validity of PGA is satisfied when scores correlate with phenomena subsequently influenced by disease activity such as quality of life measurements (HRQoL, SF-36, FACIT-Fatigue score, Lupus Impact Tracker (LIT), LupusPRO), biomarkers levels (complement fractions, ESR, auto-antibodies), treatment variations and damage assessments (SDI).

Twenty-nine studies (25, 31, 34-39, 41, 43, 48-49, 53-55, 59, 61-64, 74-75, 81, 82, 86, 92-93, 96-97) have assessed criterion validity of PGA (table 2).

**Table 2:**
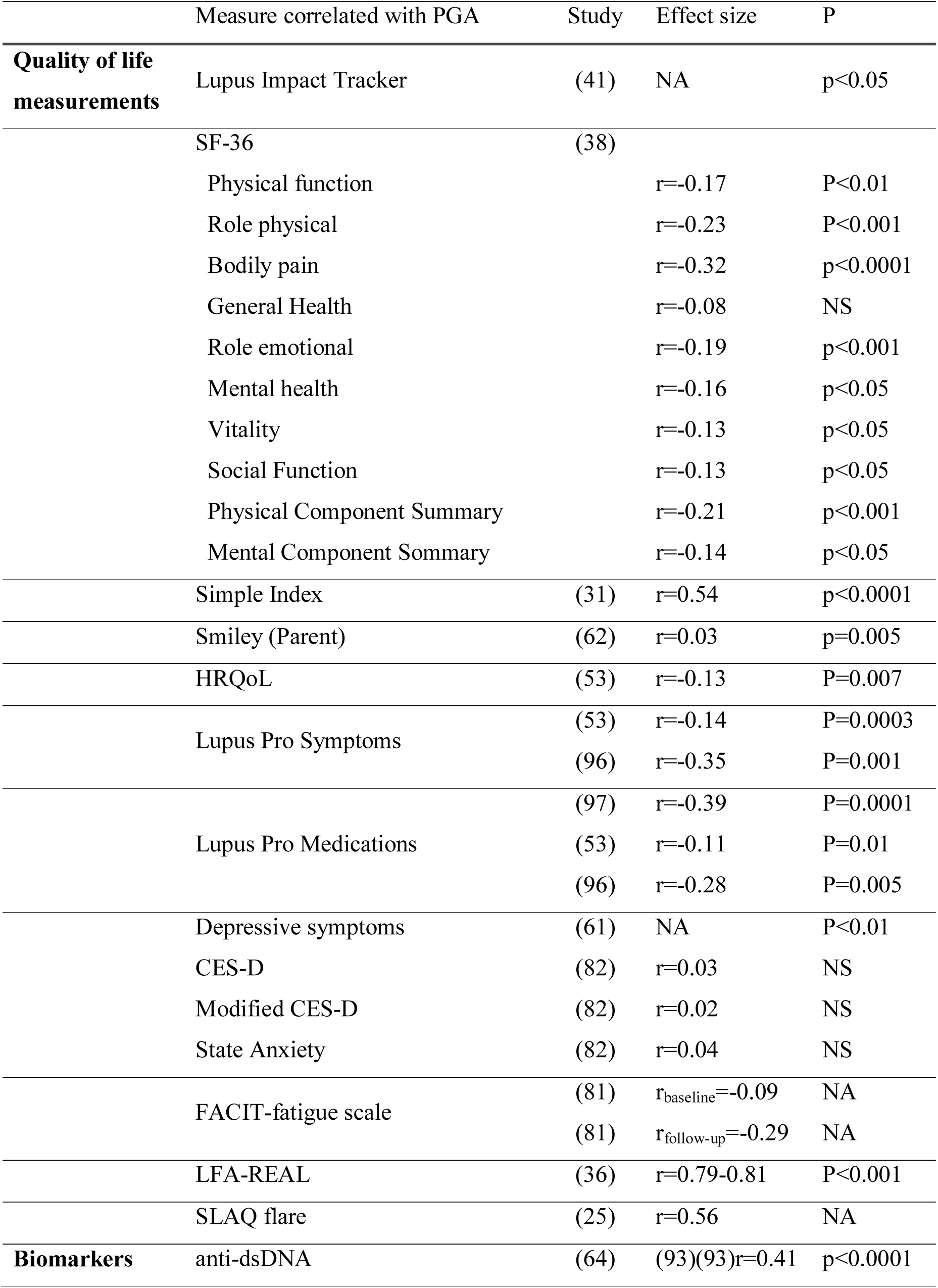
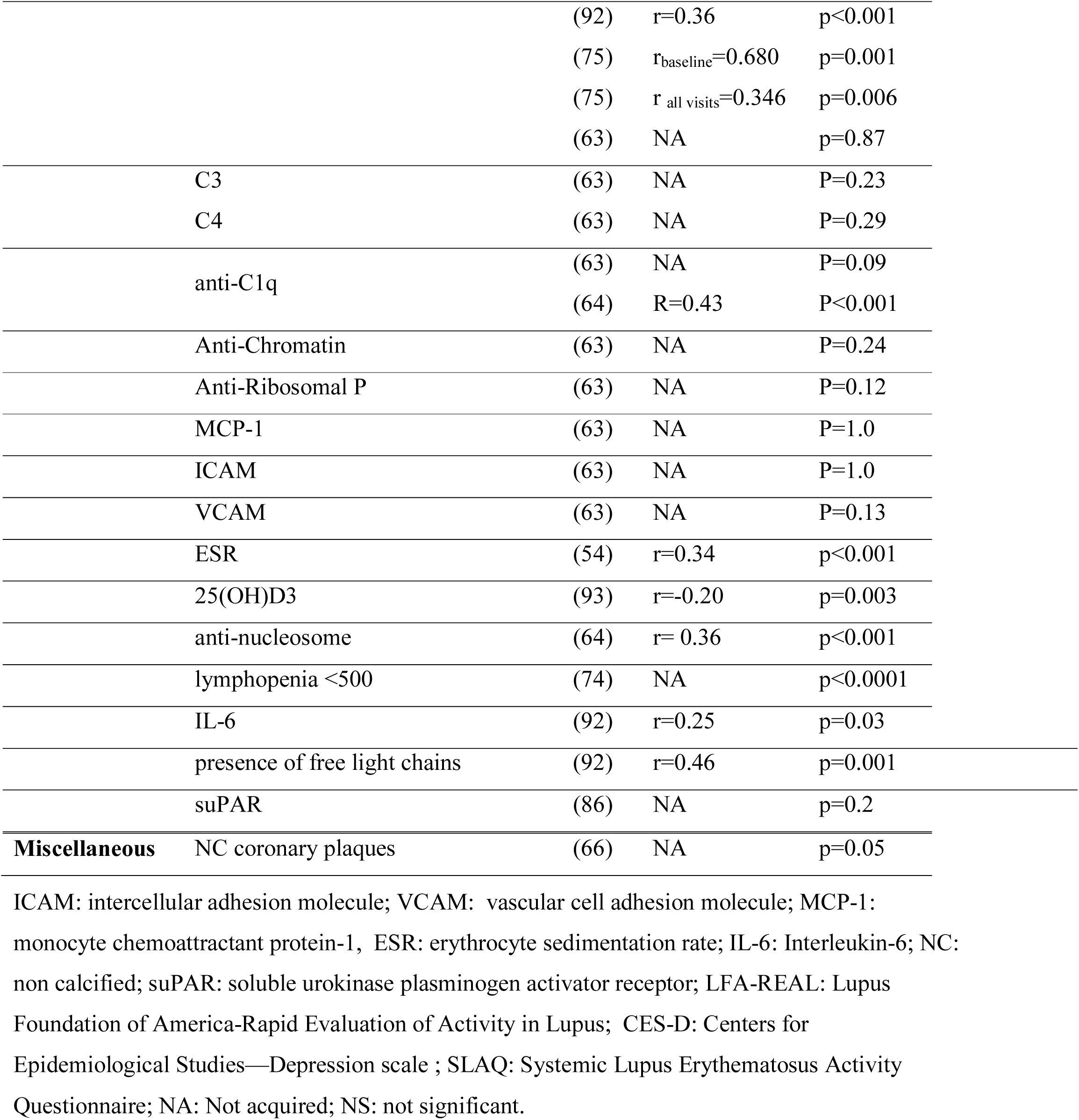
criterion validity data reporting correlation coefficients between PGA and quality of life measures, laboratory markers and miscellaneous.

Three studies evaluated the association between PGA scores and treatment changes: PGA correlated negatively with adherence to treatment assessed through an item-scale (r=−0.31, p=0.11) (34); clinically defined mild and moderate flares had a higher disease activity by PGA (P<0.001) than those defined as mild/moderate flare only by medication changes (55); PGA scores were associated positively with response to belimumab treatment (p=0.039) (43).

A PGA>1 was predictive of polymorphic light cutaneous eruption (p=0.02) (59) and correlated negatively with LLDAS attainment (37). A PGA≥2 correlated with risk of pregnancy loss (29% vs 8%, p=0.005) (49). No study has evaluated the correlation of PGA with damage measures.

#### 3.2.2 Discrimination

Discrimination refers to whether the score (PGA-VAS) differentiates between situations of interest (18): discrimination of PGA measures the ability of the PGA-VAS to report a consistent score where no change in disease activity has occurred (reliability) and to detect change when a change in disease activity has occurred (sensitivity to change or responsiveness).

##### Reliability

Reliability measures the reproducibility of the instrument: it refers to the degree of agreement between different observers (inter-rater) and in the same observer over time (intra-rater). The quantification of reliability is expressed by a correlation coefficient. An acceptable reliability is indicated by values of ICC or weighted Kappa (k) in excess of 0.60 and a good reliability is in excess of 0.85 (20).

Inter-rater reliability (Inter-RR) of PGA is the ability to provide consistent scores in a stable population between two or more physicians who evaluate disease activity of the same patient. Inter-RR was assessed in 7 studies (4, 10-11, 36, 65, 68, 94), showing values ranging from 0.67 (68) to 0.96 (94). FitzGerald and Grossman (10) found a good Inter-RR in retrospective assessment of PGA (k=0.79).

A difference between Inter-RR of PGA assessed by an untrained physician (ICC=0.5-0.63) or a trained investigator (ICC=0.79-0.81) was found (36).

The four-point PGA (0: no flare, 1 mild, 2 moderate, 3 severe) showed the lowest Inter-RR in assessing flare (ICC=0.18), compared to that of BILAG-2004 (ICC=0.54) and SFI (ICC=0.21) (65).

Intra-rater Reliability (Intra-RR) is the ability to provide consistent scores in a stable population by the same assessor over time. It estimates how similar a given patient scores were at the two visits. PGA Intra-RR was assessed in 3 studies (10, 68, 94), ranging from 0.55 (68) to 0.88 (10).

##### Responsiveness

Responsiveness, or sensitivity to change, is the usefulness of a test to detect minimum clinically important differences (MCID) (20, 109). Responsiveness of PGA is the ability to detect worthwhile variations in disease activity over time, measuring worsening or improvements in SLE disease status.

The assessment of PGA responsiveness was performed in 10 studies (4, 23, 50, 58, 77-79, 81, 83-84) using different methods (110). Only in one study (4), PGA sensitivity was assessed comparing the change with an anchor (109), represented by the treatment sensitive index (TSI): PGA sensitivity resulted comprised between that of the BILAG (highest sensitivity) and SLEDAI (lowest sensitivity). More frequently, responsiveness was assessed correlating changes in PGA with changes in other scores (23, 50, 58, 77, 78, 81, 83), finding a significative correlation with variations in SLEDAI (r ranging from 0.39-0.66) (23, 77-78), SLAM (0.61) (77), LAI (0.56) (77), ptGA (0.37) (77), SRI-50 (0.48) (78) and ESR (p<0.0001) (58), but not with C3, C4, circulating immunocomplexes and prednisone dose (77). One study showed a significative ability of the PGA in distinguishing between patients (p<0.0001), observers (p<0.0001), but not between visits (79). Ward et al (84) expressed the sensitivity in PGA scoring with the standardized response mean (SRM), demonstrating a very large effect size (ES=2.23) (110).

PGA responsiveness was used to assess flare (9): PGA was identified as the “gold standard” to rate the exacerbation of lupus activity (21, 67, 88), preliminarily defined by a change of ≥1.0 in a 0-3 VAS occurred since the last visit. To discriminate between severity of flares, PGA was incorporated in a composite index: the Selena Flare Index (10) (table 1). Several definitions of MCID were retrieved: in SRI-4 a significant worsening was defined as an increase of >10% on the PGA-VAS (111) corresponding to ≥0.3-points from baseline; Touma et al (80) considered worsening any increase in PGA from baseline; in the epratuzumab trial (87) a significant improvement was a 20% decrease in PGA score evaluated after 12 months of treatment.

In a post-hoc analysis of phase 3 Belimumab trials, improvements and no worsening in PGA were greater among SRI responders versus SRI non-responders (p<0.001) (52). Otherwise, a validation study of the SRI for juvenile SLE (60) showed that the exclusion of the BILAG or PGA from the SRI did not change the accuracy of the SRI in detecting improvement.

#### 3.2.3 FEASIBILITY

Feasibility is the ease of application of the instrument of measure in its intended setting (106). Feasibility refers not to the quality of the outcome measure, but to aspects such as completion time, cost of an instrument, equipment, type and ease of administration. No study has evaluated the feasibility of the PGA in SLE to date.

## 4. DISCUSSION

The assessment of disease activity in SLE is particularly challenging. In this systematic review, we have analysed the measurement properties of the PGA, including the validity, reliability, responsiveness and feasibility.

In support of its face validity, the PGA was used to define disease activity score in all the 91 studies retrieved by the literature search, having a role as outcome measure as well as comparator to assess the validity of other indices. Nevertheless, despite the fact that the PGA was considered the reference in 39 studies involving other indices, it was used as the sole instrument in only 2 of them. This suggests that the role of the PGA is limited for disease activity assessment when used as single instrument. It is therefore desirable to use the PGA along with other tools (typically the SLEDAI) or to include PGA in a composite index (e.g., SFI, LLDAS, SRI, DORIS remission criteria) (3, 5, 10, 13-15, 104).

However, the PGA allows for the measurement of disease activity in a global way (content validity). It does not provide a predefined or limited list of disease manifestations or organ systems, therefore allowing to capture all the heterogeneous aspects of SLE disease activity. PGA is usually reported by experts as allowing an exhaustive coverage of the concept of disease activity in SLE (20, 108). Thanks to this feature, the PGA was included in composite indices with the aim to rate manifestations not included in glossary-based instruments such as the SLEDAI and BILAG (3) or for which a threshold has been defined (cytopenia).

It is noteworthy that the PGA correlates with several other instruments which measure disease activity. Construct validity is shown by the good correlation observed with the SLEDAI, BILAG, LAI and ECLAM (10, 23-24, 26, 28-29, 35, 54, 68, 76, 84, 99).

The PGA also showed a good predictive validity as it correlated significatively with measures of future outcomes, such as quality of life or laboratory exams but no study has currently evaluated its correlation with measures of damage.

Of note, the literature search revealed heterogeneous definitions of physician assessment of disease activity other than PGA (MD global (4, 70, 73), physician overall assessment [PHYOA] (85)). Different scores and lengths of visual evaluation were employed: the first was the 0-10 VAS suggested by Liang et al (8) and adopted in childhood SLE; the most common tool (the 0-3 VAS), was developed (68) to capture the concept of flare, and measured on 3 cm VAS in SRI (3) and 10 cm VAS in SFI (10, 104), but other scores (0-2, 0-4, 0-5, 0-7) (11, 53, 78, 80, 86-87) and lengths (8 cm, 15 cm) (10, 82-84) have also been used. Currently, no agreement has been reached upon which scale should be used: if a pointed scale with anchored values (0, 1, 2, 3) or a centimetric scale with all values comprised between 0.0 and 3.0. Moreover, there is an uncertainty as to whether the best timing of assessment is prior or after reviewing laboratory exams (26). The lack of standardized scoring, as well as the subjectivity of the physician judgments, can be an important source of heterogeneity, especially in trials.

Five studies have demonstrated good ICC values for reliability (all higher than 0.60 and ranging up to 0.97). Reliability was excellent when scored through a pointed scale, as the Likert scale which was anchored in unit numbers from 0 (not active) to 7 (most active) (Inter-RR ICC: 0.96; Intra-RR ICC: 0.88) (80) but lower when assessed through a centimetric VAS scale using values comprised between 0.0 and 3.0 (Inter-RR ICC: 0.67; Intra-RR ICC: 0.55) (68). This important heterogeneity in the anchoring of the PGA prevented us from performing a metanalysis of reliability data.

Physician training is very important. Even though the PGA showed optimal reliability, a very low Inter-rater reliability for flare using the PGA (ICC=0.18) was found in a single study (65), compared to that of the BILAG (ICC=0.54) or SFI (ICC=0.21). According to the authors, this difference was probably due to the greater familiarity of the physicians with the BILAG 2004 index. Moreover, a difference between Inter-RR of PGA assessed by an untrained physician (0.5-0.63) and a trained investigator (0.79-0.81) was found, suggesting the need for a PGA-scoring training or a standardization (36).

Responsiveness of the PGA was assessed through different methods (109, 111) showing a high sensitivity for detecting clinical variations (84). Changes in PGA correlated with changes of other disease activity indices (SLEDAI, SLAM, LAI, ptGA), laboratory exams (ESR), PROs (LIT) (23, 50, 58, 77-78, 81, 83) and response to treatment (4). These results enabled its use as a gold standard for assessing flare and defining flare severity in several studies (21, 67, 88).

No data were found regarding feasibility of the PGA. In most studies, the PGA was assessed by a rheumatologist experienced in SLE care or research and, as already stated, the ICC reliability was different between an untrained physician and a trained investigator (36).

Finally, the PGA enables the measurement of disease activity globally compared to a glossary-based index. At the same time, a global judgment does not allow the detailed capture of specific organ manifestations of the disease. Altogether, the PGA should ideally be used in a composite index or as part of a disease status definition.

**In conclusion**, the PGA was demonstrated to be a valid, responsive and feasible instrument, while its reliability was strongly impacted by the scale adopted, suggesting the major need for a standardization in its scoring.

## Data Availability

Data are publicly available in the literature (non proprietary data)

## Acknowledgment

The authors wish to thank Ms. Sylvie Thuong for her invaluable assistance in the preparation of the manuscript.

